# Prevalence and factors influencing contraceptive uptake among in school adolescent girls aged 13-19 years in selected secondary schools in Lusaka district

**DOI:** 10.1101/2024.11.05.24316787

**Authors:** Jennise Mubanga, Alice Ngoma Hazemba, Phoebe Mwembya

## Abstract

**Background:** Low contraceptive uptake among in school adolescent girls continues perpetuating poor sexual and reproductive health outcomes. Adolescent pregnancy has adverse maternal complications and compromise their social-economic potential. Studies have shown high prevalence of adolescent pregnancy due to low contraceptive uptake, despite availability of effective preventive measures. Adolescents still have many unmet contraceptive needs. Therefore, contraceptive use among in school adolescent girls needs more attention.

**Methods:** A cross-sectional design was conducted among 383 in school adolescent girls aged 13-19. Primary data was collected and analyzed using STATA version 13.0. Characteristics of respondents were analyzed and presented in Frequency and Percentages. Applied chi-square and fishers exact tests to measure association between response variable and explanatory variables. Applied logistic regression to determine odds of factors influencing contraceptive uptake. Used investigator led best fit model to select best predictors of contraceptive uptake with statistically significant p-value (<0.05) and 95% confidence-interval.

**Results:** Study showed majority 70% of adolescent girls never used contraceptives while, 20.6% ever used. Odds of contraceptive uptake increased among adolescents with both moderate and high knowledge levels (AOR:7.38, 95% CI:1.16-46.87, p =0.034) and (AOR:66.73, 95% CI3.84-1161.01, p = 0.004), having moderate and easy access (AOR:15.98, 95% CI:5.01-50.95, p<0.000) and (AOR:893.57, 95% CI:38.84-20560.12, p<0.000). Confidentiality showed moderately and extremely confidential (AOR:6.84, 95%CI: 1.96-23.85, p =0.003) and (AOR:124.60, 95%CI:6.07-2558.39, p =0.002). Also, having satisfactory and excellent information about sexual and reproductive health showed high odds (AOR:12.53, 95% CI:3.86-40.67, p<0.000) and (AOR:25.13, 95% CI:.3.34-188.95, p =0.000 respectively.

**Conclusion:** Findings highlighted low contraceptive prevalence with factors influencing contraceptive uptake such as age, knowledge, access, confidentiality and adequate sexual and reproductive health information. Therefore, strengthening collaboration between education and healthcare systems in meeting sexual and reproductive health needs would significantly reduce teen pregnancies and promote their socio-economic development.

## Introduction

“Knowledge is power. Equipping adolescents with accurate, accessible information empowers them to make informed choices about their health and future. Contraceptive uptake among adolescents is essential to prevent unwanted pregnancies and the associated health risks. Adolescence is a crucial transition from childhood to adulthood, a period marked with specific health and developmental needs and rights. As many adolescents begin to engage in sexual activity during this stage, providing them with adequate sexual and reproductive health information is vital to support safe, responsible decision-making”.

Globally, sexual and reproductive health (SRH) needs of adolescents have remained largely unmet with about 20 million adolescent girls aged 15-19 in need of modern contraceptive methods ^1^. According to a report published in 2016 on the “Costs and Benefits of Meeting Contraceptive Needs of Adolescents,” about 21 million girls aged 15-19 become pregnant yearly in developing countries, about 12 million give birth and about 10 million pregnancies are unwanted ^2, 3, 4^.

Adolescent pregnancy and childbirth complications are the leading causes of maternal and neonatal deaths ^1, 5^. Although, global birth rate reduced to 44 births per 1,000 girls from 2015 to 2020 and Sustainable Development Goal (SDG) 3 has emphasized the need to ensure healthy lives, promote wellbeing for all at all ages, reduce maternal mortality to less than 70 per 100,000 live births by 2030 and grant universal access to sexual and reproductive care, family planning and education ^6^. This is far from being achieved as low and middle income Countries (LIMCs) are still lagging behind compared to industrialized nations. Many studies have focused on contraceptive needs of married women of reproductive age 15-49 leaving unmarried and younger adolescent 10-14 with high unmet needs for contraceptive. Also, very few studies have been done on contraceptive needs of in school adolescent girls where majority of this population is found. Therefore, there is need to develop and implement effective sexual and reproductive health educational programs within schools and healthcare facilities as well as reinforce existing ones. The programs should highlight the importance of reproductive health, provide easy access to contraceptive and relevant sexual and reproductive health information. Failure to take immediate action would continue perpetuating the cycle of poor reproductive health outcomes among adolescents.

## Materials and Methods

### Study design

The study used a quantitative cross sectional design which involved participation of in school adolescent girls aged 13-19 from three selected secondary schools in Lusaka district.

### Study population

A total of 383 participants was included in the study and a non-probability convenient sampling method was used to select study participants.

#### Data collection and methods

Primary date was collected by administering structured questionnaires to participants. Data was collected by the researcher.

#### Variables

The study included variables that are categorized into constructs: Socio-demographic factors (age, education and knowledge), education system factors (comprehensive sex education, adequate sexual and reproductive health information and teacher support) and Health system factors (access, availability and confidentiality).

#### Primary outcome

The primary outcome was contraceptive uptake. From the questionnaire, the question was phrased as “Have you ever used any method of contraceptive before? those who responded YES were categorized as ever used contraceptives and those who responded NO were categorized as never used contraceptives.

#### Independent variables

These variables were categorized as follows: *Socio-demographic variables*: age, education and knowledge

#### Education system variables

Comprehensive sex education, adequate sexual and reproductive health information and teacher support

#### Health system variables

Accessibility, availability and confidentiality.

### Data analysis

Data was analyzed using Stata version 13.0. The characteristics of respondents were analyzed and presented using Frequency and Percentages. The study applied Chi-square and fishers exact tests to measure the association between the response variable and explanatory variables. Logistic regression was applied to determine the odds of factors influencing uptake of contraceptives among in school adolescent girls. An investigator led best fit model was used to select the best predictors of contraceptive uptake with statistically significant p-value less than (<0.05) and 95% confidence interval.

### Ethical Approval

Ethical approval was sought from the University of Zambia biomedical research committee (UNZABREC), National Health Research Authority (NHRA), Ministry of Health and District Education Board Secretary for permission to conduct the study. Due to sensitivity of the study with regards to religion, culture and socio-political controversy parental consent and assent were sought from all participants and high level of confidentiality was maintained.

## Results

### Descriptive statistics

Table 1 The study comprised 383 total participants with vast majority in the age range 15-17 (52.48%) with the lowest age 18-19 (15.93%). Regarding education level, senior secondary level showed 63.71% out-numbering their junior counterparts 36.29%. Knowledge level, showed majority with low knowledge 50.65% while, a smallest subset showed high knowledge level 8.88%. Accessibility of services showed vast majority 63.45% found it difficult to access while, minority 11.75% indicated easy access. Furthermore, participant’s perspective on the availability of services showed majority 73.11% disagreed while, the smallest subset 9.40% agreed to availability of contraceptive services. Regarding confidentiality, majority 61.88% found services slightly confidential and minority 9.66% indicated extremely confidential. Majority 66.06% expressed having learnt comprehensive sex education while, 33.94% expressed not having learnt. In terms of having adequate sexual and reproductive health (SRH) information, majority 69.71% indicated having inadequate information while, minority 12.01% expressed having excellent information. Regarding teacher support, vast majority 83.29% expressed a neutral support, and the lowest 3.39% agreed to teacher support.

**Table 1:** The characteristics of study participants aged 13-19 (n=383)

| **Basic variables** | **Frequencies** | **Percentage (%)** |
| --- | --- | --- |
| **Age** |  |  |
| 13-14 | 121 | 31.59 |
| 15-17 | 201 | 52.48 |
| 18-19 | 61 | 15.93 |
| **Education level** |  |  |
| Junior secondary level | 139 | 36.29 |
| Senior secondary level | 244 | 63.71 |
| **Knowledge level** |  |  |
| Low | 194 | 50.65 |
| Moderate | 155 | 40.47 |
| High | 34 | 8.88 |
| **Accessibility** |  |  |
| Difficulty | 243 | 63.45 |
| Neutral | 95 | 24.80 |
| Easy | 45 | 11.75 |
| **Availability** |  |  |
| Disagree | 280 | 73.11 |
| Neutral | 67 | 17.49 |
| Agree | 36 | 9.40 |
| **Confidentiality** |  |  |
| Slightly confidential | 237 | 61.88 |
| Moderately confidential | 109 | 28.46 |
| Extremely confidential | 37 | 9.66 |
| **Comprehensive-sex education** |  |  |
| No | 130 | 33.94 |
| Yes | 253 | 66.06 |
| **Adequate-SRH-information** |  |  |
| Inadequate | 267 | 69.71 |
| Satisfactory | 70 | 18.28 |
| Excellent | 46 | 12.01 |
| **Teacher support** |  |  |
| Disagree | 51 | 13.32 |
| Neutral | 319 | 83.29 |
| Agree | 13 | 3.39 |

Source: Field Data (2023)

### Prevalence of contraceptive uptake

Figure 1. : Contraceptive prevalence indicates 20.63% of adolescent ever used, while majority 79.37% never used.

**Figure 1.**
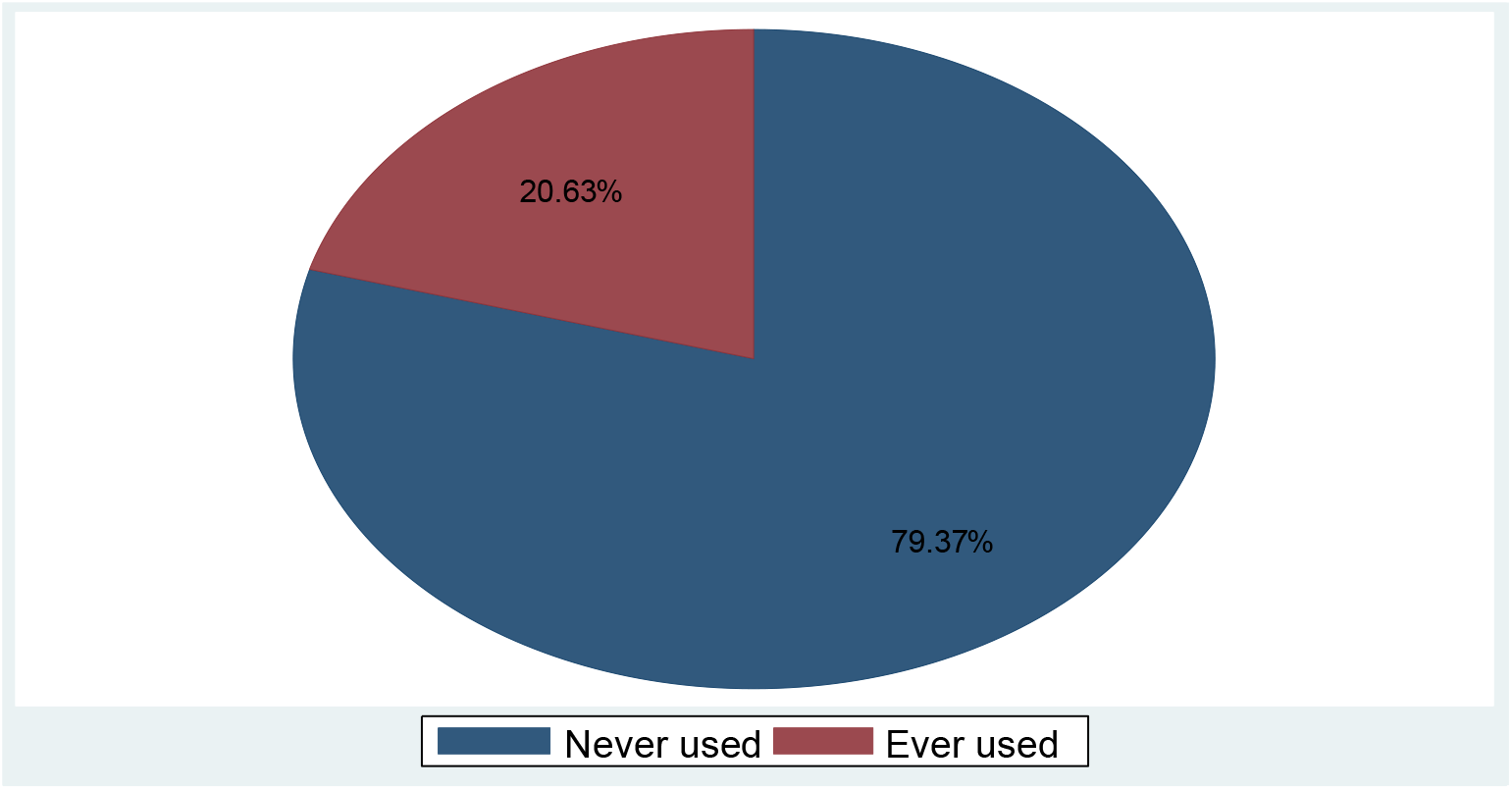
Prevalence of contraceptive uptake among in school adolescent girls age 13-19 (n=383)

### Factors associated with contraceptive uptake

Table 2: shows chi square and fishers exact test of association between each explanatory variables and response variable. All variables showed association with contraceptive uptake with 95% CI, p <0.000.

**Table 2:**
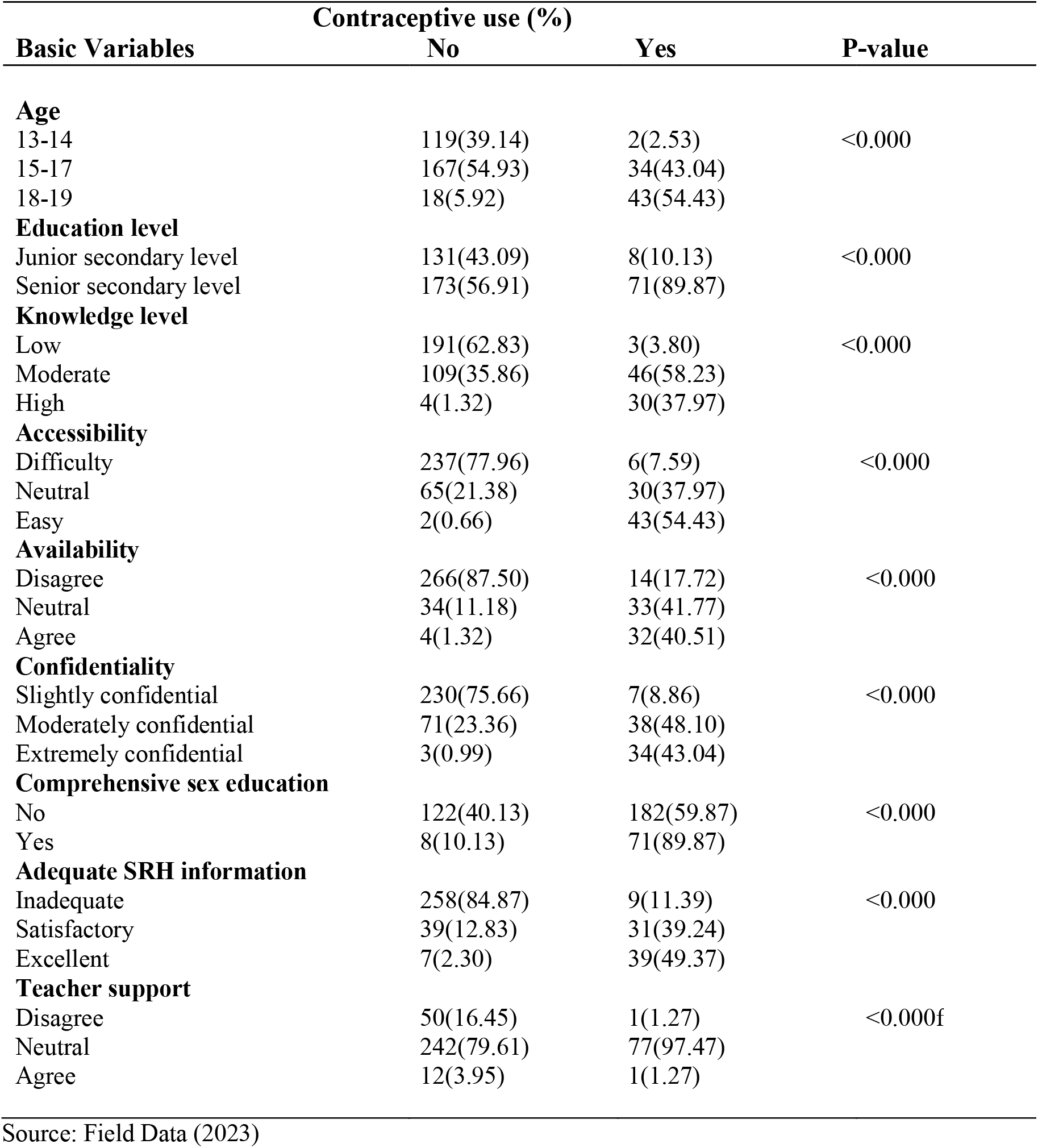
Factors associated with contraceptive uptake among in school adolescent girls aged 13-19 (n-383)

| Basic Variables | Contraceptive use (%) |  | P-value |
| --- | --- | --- | --- |
|  | No | Yes |  |
| **Age** |  |  |  |
| 13-14 | 119(39.14) | 2(2.53) | <0.000 |
| 15-17 | 167(54.93) | 34(43.04) |  |
| 18-19 | 18(5.92) | 43(54.43) |  |
| **Education level** |  |  |  |
| Junior secondary level | 131(43.09) | 8(10.13) | <0.000 |
| Senior secondary level | 173(56.91) | 71(89.87) |  |
| **Knowledge level** |  |  |  |
| Low | 191(62.83) | 3(3.80) | <0.000 |
| Moderate | 109(35.86) | 46(58.23) |  |
| High | 4(1.32) | 30(37.97) |  |
| **Accessibility** |  |  |  |
| Difficulty | 237(77.96) | 6(7.59) | <0.000 |
| Neutral | 65(21.38) | 30(37.97) |  |
| Easy | 2(0.66) | 43(54.43) |  |
| **Availability** |  |  |  |
| Disagree | 266(87.50) | 14(17.72) | <0.000 |
| Neutral | 34(11.18) | 33(41.77) |  |
| Agree | 4(1.32) | 32(40.51) |  |
| **Confidentiality** |  |  |  |
| Slightly confidential | 230(75.66) | 7(8.86) | <0.000 |
| Moderately confidential | 71(23.36) | 38(48.10) |  |
| Extremely confidential | 3(0.99) | 34(43.04) |  |
| **Comprehensive sex education** |  |  |  |
| No | 122(40.13) | 182(59.87) | <0.000 |
| Yes | 8(10.13) | 71(89.87) |  |
| **Adequate SRH information** |  |  |  |
| Inadequate | 258(84.87) | 9(11.39) | <0.000 |
| Satisfactory | 39(12.83) | 31(39.24) |  |
| Excellent | 7(2.30) | 39(49.37) |  |
| **Teacher support** |  |  |  |
| Disagree | 50(16.45) | 1(1.27) | <0.000f |
| Neutral | 242(79.61) | 77(97.47) |  |
| Agree | 12(3.95) | 1(1.27) |  |

Source: Field Data (2023)

### Factors associated with contraceptive uptake (bivariable and multivariable)

Table 3: The results from bivariate logistic regression analysis show factors significantly associated with contraceptive uptake among in school adolescent girls: age, education, knowledge, accessibility, availability, confidentiality, comprehensive sex education, adequate SRH information and teacher support. To obtain adjusted estimate for contraceptive uptake multivariable logistic regression analysis was used. Adolescent girls aged 15-17 showed no significant difference compared to the reference (13-14) with odds (OR:0.32 CI:0.04-2.79, p = 0.304). While 18-19 years old showed significant decreased odds (: 0.00 0.00-0.05, p = 0.001). High knowledge and moderate knowledge level both had increased odds (OR:66.73 CI:3.84-1161.01, p = 0.004) and (OR:7.38 CI:1.16-46.87, p = 0.034) compared to the reference group (low). Neutral response to accessing services was significantly associated with contraceptive uptake (OR:15.98 CI:5.01-50.95, p<0.000). Easy access responses also showed increased odds of (OR: 893.57 CI:38.84-20560.12, p<0.000) compared to the reference category (difficult). Confidentiality of contraceptive services showed both moderately confidential and extremely confidential perceptions were significantly associated with increased contraceptive uptake (OR:6.84 CI:1.96-23.85, P =0.003) and (OR: 124.60(6.072558.39, p =0.002) compared to reference category (slightly confidential) respectively. Adequate sexual and reproductive health information (SRH) showed both satisfactory (OR:12.52 CI:3.86-40.67, p<0.000) and excellent (OR:25.13 CI:3.34-188.95, p =0.002) sexual and reproductive information were associated with increased contraceptive uptake compared to the reference category (inadequate) respectively.

**Table 3:**
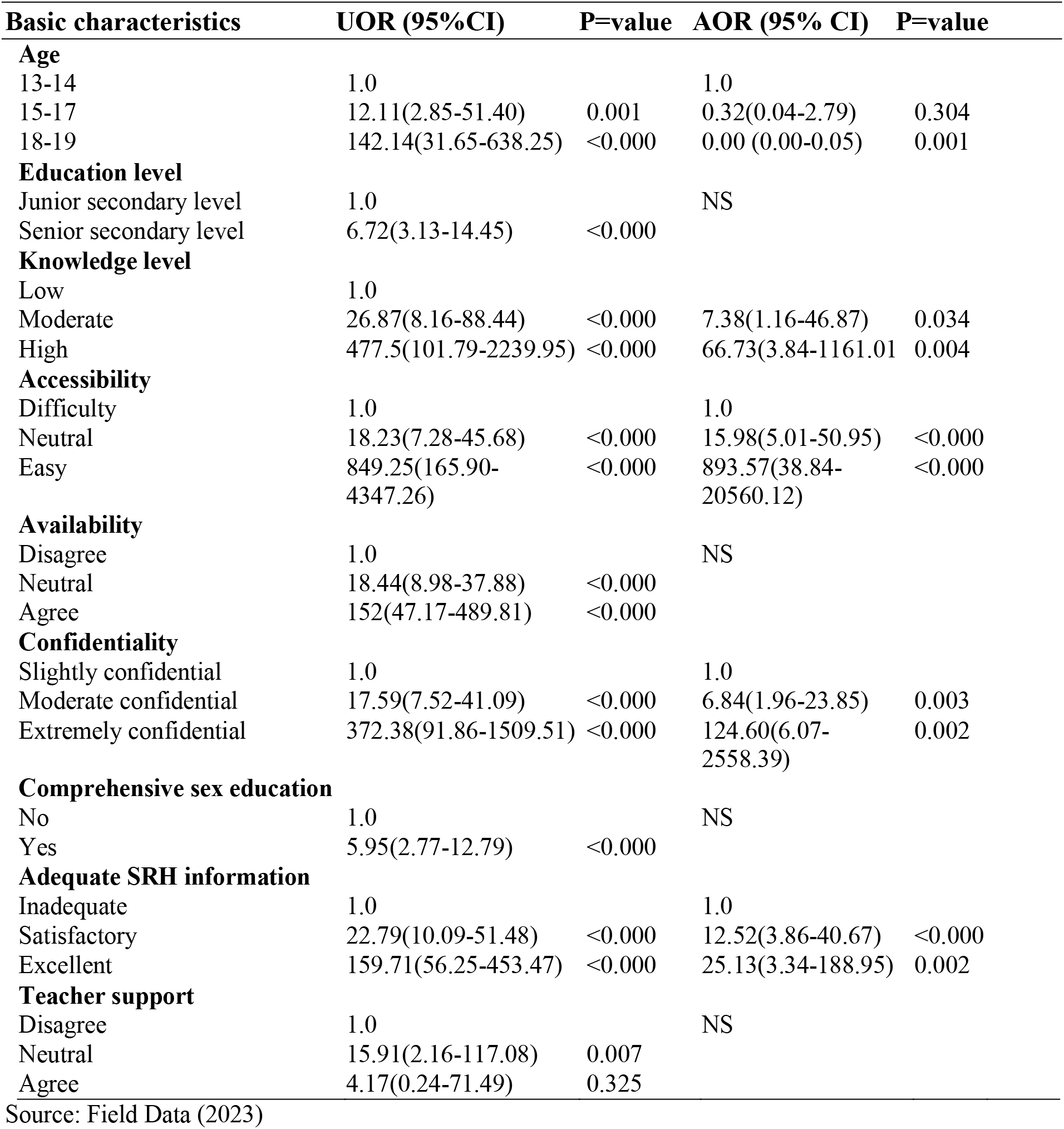
Factors associated with contraceptive uptake among in school adolescent girls age 13-19 (n=383)

## Discussion

The study aimed to assess prevalence of contraceptive uptake and determine factors influencing contraceptive use among in school adolescent girls. In this study, 20.63% adolescent girls aged 13-19 were reported ever used contraceptives. Age, knowledge, access, confidentiality and adequate sexual and reproductive health information were significantly associated with contraceptive uptake.

The current study found prevalence of contraceptive uptake among in school adolescent girls aged 13-19 to be low. Only a fifth of the population used contraceptives. The findings of this study are however slightly higher than those in the Zambia National Demographic Health Survey ^7^ among adolescents aged 15-19 in comparison to the current study. Meanwhile, the context of sub-Sahara Africa showed comparatively higher contraceptive prevalence. In contrast, global estimates in 2019 showed lower prevalence, with proportion of adolescent women aged (15–19 years) who were using modern methods of contraception. This shows that uptake of contraceptive among adolescent girls is very low with high unmet needs for contraceptives. The reasons for the difference in contraceptive prevalence rates among these studies may be attributed to methodological differences. The current study used a broader age range 13-19 whereas ZDHS, Sub-Sahara Africa and global/regional estimates used a narrower age range of 15-19. This study collected date by administering questionnaires to participants who were in school at the time of the study. Differing from the current study, the ZDHS, Sub-Sahara Africa and global/regional studies used a comprehensive dataset of family planning indicators among women aged 15-19 who were both in and out of school. In the case of global/regional survey, a Bayesian hierarchical model with country specific annual trends was used to estimate contraceptive prevalence and unmet needs for family planning for 185 countries^8^. The exposure of in school adolescents to comprehensive sexuality education and sexual and reproductive health information would have contributed to the slight increase in prevalence rate in the current study. Also, the introduction of free education in secondary schools by the government in 2021 facilitated for high enrolment of girls, this too may have contributed to a modest increase of contraceptive prevalence rate (CPR). However, this study reported low prevalence compared to the study done in Kenya, with slightly high contraceptive prevalence among adolescent girls aged 15-19 in secondary schools who were sexually active ^9^. But, the study left the equally important group of adolescents those below 15 years as they have contraceptive needs as well since some start sexual activities at a tender age. Different from the current study, a study in Malawi showed lower prevalence rate among unmarried sexually active adolescent girls ^10^. In another study done in Nigeria on contraceptive use among adolescent girls reported that modern contraception use is very low ^11^. Also, a study conducted in Zambia ^12^ showed that contraceptive use remains low in 1996 to 2013/14, reflecting very minimal increase over 18 years. Although different studies showed varying contraceptive prevalence rates, they all fall below fifty percent. Suggesting low contraceptive prevalence and high unmet needs of contraceptive among adolescent girls thus, predisposing them to poor sexual and reductive health outcomes.

### Demographic factors influencing contraceptive uptake among in school adolescent girls

In this study, age showed a difference in contraceptive use among adolescent girls across different age categories, with 13-14 age group being the lowest while 18-19 age group being the highest. However, 18-19 age group exhibited a significant decrease in contraceptive use. In contrast, a study done in Zambia ^2^ showed that over the period 1996 to 2013/14, contraceptive use increased among 19-year-old and lowest in 15-year-old. Another, study conducted in kiambu county, Kenya ^13^ indicated that uptake of contraceptives increased in tandem with age. Adolescent girls who were 18 years of age were more likely to use contraceptives as compared to their counterparts of less age. The trend in contraceptive use with age groups can be attributed to age related restriction. A qualitative study done in Nigeria ^14, 15^ showed that age related restriction of adolescent having access to contraceptives contributed to adolescents less than 18 years not utilize contraception services. Another study in Nigeria ^16^ points out that adolescent age influence use of contraceptive methods. Adolescent girls below 18 years find it difficult to access and use contraceptives. Several studies have shown that age is a hindrance to contraceptive uptake. The variation observed in these studies can be attributed to a number of reasons. The current study population consisted only of school going adolescent girl who could have had different level of knowledge, access and cultural factors affecting their contraceptive use compared to the general population. Also, uneven age distribution across all age categories, 18-19 age group had the smallest sample size proportion of participants resulting in more variability and less statistical power to detect effect.

The study showed majority had low contraceptive knowledge level with moderate and high knowledge level exhibiting significantly high odds ratio, suggesting a notable association with increased contraceptive uptake among adolescents. Similarly, a study done in Rwanda ^17^ showed that majority of secondary school female adolescents had inadequate knowledge on contraceptives. Another study conducted in central region of Sudan indicated that less than a half participants had adequate knowledge ^18^. However, different results were found in the study done in Nigeria which showed that the majority of participants had adequate knowledge about contraceptives. This is probably due to the difference in cultures among these countries. A study done in Malawi ^19^ showed high contraceptive knowledge level among adolescent girls at Tsangano community. It indicated that respondents with knowledge were 4 times more likely to use modern contraceptives than their counterparts without knowledge. Another, study conducted in Kenya^3^ indicated that among sexually active adolescents, having knowledge on contraceptives increases by three times the likelihood to use contraceptive. Having knowledge gives one power to make informed decision. This study also agrees with the findings by WHO which states that knowledge about contraceptive is an effective way of reducing teenage pregnancies as adolescents who have the knowledge are able to use modern contraceptives ^20^.

### Healthcare system factors influencing contraceptive uptake among in school adolescent girls

The majority of participants indicated that access to contraceptive services was difficult, neutral and easy responses showing significantly high odds, indicating significant association with increased uptake of contraceptive. This aligns with the study done in Kenya ^4^ which showed that having access to contraceptive was significantly associated with increased uptake of contraceptives. The study revealed that uptake of contraceptive increases with increase in access as those who indicated having access were more likely to use contraceptives services than those who did not.

The study showed majority participants expressed slightly confidential to contraceptive services and those who indicated moderately and extremely confidential had high odds of contraceptive use. In this study, extremely confidential showed very high association of contraceptive uptake. Those who expressed moderately confidential were six times more likely to use contraceptive. A qualitative study in Ethiopia reported that adolescent girls who were not using modern contraception complained lack of confidentiality as the reason for their failure to using the services^21^. These studies suggest that confidentiality of contraceptive services is an influencing factor to uptake.

### Education system factors influencing contraceptive uptake among in school adolescent girls

The study showed that majority participants had inadequate information on sexual and reproductive health while, minority indicated having satisfactory and excellent information. Both satisfactory and excellent showed significantly high association with contraceptive uptake. Signifying increase in SRH information increases contraceptive uptake. A study conducted in Saudi Arabia to determine sexual health understanding among teenage girls in schools, revealed that the determinant for poor sexual health education in adolescents was the ineffectiveness of a school curriculum on sexual health^22^. Other studies conducted in Nepal and Iran, showed that many young people, particularly unmarried girls, have inadequate sexual health information and are less likely to use contraceptives ^23, 24^. Therefore, immeasurable basic understanding necessary for safer sexual behavior is missed. Their findings also indicated that secondary students were exposed to comprehensive sex education programmes that had greater knowledge and were intended to make teenagers refuse sex and decrease the frequency of sex but did not emphasize on contraceptive use and its importance. A research at Guttmacher Institute in New York, indicated that most unplanned pregnancies experienced by teenage girls arise among those who use no birth control measures ^5^. This shows that although sex education is integrated into the school curriculum majority of adolescent girls have inadequate information on sexual and reproductive health (SRH). On the other hand, having knowledge of SRH evidently increases contraceptive uptake.

### Strengthens and limitations

The study provided primary information concerning contraceptive uptake among in school adolescent girls. This would help future researchers, health system and education system to be aware of the many challenges adolescent girls face with regards to sexual and reproductive health outcomes. This would also help in formulating well focused intervention programs that would promote contraceptive use and reduce problems of adolescent pregnancies.

The study faced challenges with data collection from participants due to scheduling conflicts with their classes, making it difficult to include as many girls within a short period of time. But somehow we managed to reach our sample size.

Literature on contraceptive uptake among in school adolescent girl was limited, many studies predominantly centered around adolescent in the general population rather than specifically addressing those in a school setting.

The study focused on school going adolescent girls among three selected secondary schools in Lusaka District and therefore, did not reference to the total population of adolescents given that a large proportion of adolescent girls were out of school. The study also applied descriptive study design that was cross sectional in nature and data was collected at one point in time thus, limiting the generalization.

## Conclusion

From the findings, this study discovered that uptake of contraceptive continues to be very low among adolescents. Despite contraceptive being one of the most effective pregnancy preventive measures, uptake remains low specially in LMIC like Zambia. Many factors such as age, low knowledge, difficult in accessing services, lack of confidentiality of services and inadequacy of sexual and reproductive health information influence contraceptive uptake among adolescent girls. This emphasize the need for school system and health system to improve on the delivery of sexual and reproductive health information and services to adolescents. This will improve their sexual and reproductive health and enable them to attain their full socioeconomic potential.

## Data Availability

all data produced in the present study are available upon reasonable request to the authors

## Author contribution

JM: conceptualization, methodology, analyzed data, interpreted the findings, writing original draft, writing-reviewing and editing. AH: Guided from conceptualization to implementation and reviewing. PB: Guided from conceptualization to implementation and reviewing.

## Acknowledgement

The University of Zambia Biomedical Research Ethics Committee (UNZABREC) and National Health Research Authority (NHRA) for granting permission to carry out the research. To participants, thank you for agreeing to be part of the study. To my supervisors Dr Alice Ngoma Hazemba and Dr Phoebe Bwembya, I am grateful for your guidance throughout the study period.

## Ethical approval

The study sought ethical clearance from the University of Zambia Biomedical Research Committee (REF.No.3596-2022) and National Health Research Authority (NHRA) for permission to conduct the study. Also, due to the nature of the study written consent and assent were sought form participants.

## Footnotes

1 **Error! Bookmark not defined.**

2 ibid

3 13

4 13

5 2

